# Long-lasting neutralizing antibody responses in SARS-CoV-2 seropositive individuals are robustly boosted by immunization with the CoronaVac and BNT162b2 vaccines

**DOI:** 10.1101/2021.05.17.21257197

**Authors:** Nicolás A. Muena, Tamara García-Salum, Catalina Pardo-Roa, Eileen F. Serrano, Jorge Levican, María José Avendaño, Leonardo I. Almonacid, Gonzalo Valenzuela, Estefany Poblete, Shirin Strohmeier, Erick Salinas, Denise Haslwanter, Maria Eugenia Dieterle, Rohit K. Jangra, Kartik Chandran, Claudia González, Arnoldo Riquelme, Florian Krammer, Nicole D. Tischler, Rafael A. Medina

**Affiliations:** Laboratorio de Virología Molecular, Fundación Ciencia & Vida, Av. Zañartu 1482, Santiago, Chile; Department of Pediatric Infectious Diseases and Immunology, School of Medicine, Pontificia Universidad Católica de Chile, Santiago, Chile; Advanced Interdisciplinary Rehabilitation Register (AIRR) – COVID-19 Working Group, Faculty of Medicine, Pontificia Universidad Católica de Chile, Santiago, Chile; PhD Program in Biological Sciences, Mention in Molecular Genetics and Microbiology, Faculty of Biological Science, Pontificia Universidad Católica de Chile, Santiago, Chile; Department of Pathology, Icahn School of Medicine at Mount Sinai, New York, NY, USA; Department of Microbiology and Immunology, Albert Einstein College of Medicine, New York, NY, USA; Department of Otorhinolaryngology, School of Medicine, Pontificia Universidad Católica de Chile, Santiago, Chile; Department of Gastroenterology, School of Medicine, Pontificia Universidad Católica de Chile, Santiago, Chile; Department of Health Sciences, Faculty of Medicine, Pontificia Universidad Católica de Chile, Santiago, Chile; Department of Microbiology, Icahn School of Medicine at Mount Sinai, New York, NY, USA; Facultad de Medicina y Ciencia, Universidad San Sebastián, Santiago, Chile

**Keywords:** COVID-19, serological response, neutralizing antibody persistence, SARS-CoV-2 vaccines

## Abstract

The durability of circulating neutralizing antibody (nAb) responses to severe acute respiratory syndrome coronavirus 2 (SARS-CoV-2) infection and their boosting by vaccination remains to be defined. We show that outpatient and hospitalized SARS-CoV-2 seropositive individuals mount a robust neutralizing antibody (nAb) response that peaks at days 23 and 27 post-symptom onset, respectively. Although nAb titers remained higher in hospitalized patients, both study groups showed long-lasting nAb responses that can persist for up to 12 months after natural infection. These nAb responses in previously seropositive individuals can be significantly boosted through immunization with two doses of the CoronaVac (Sinovac) or one dose of the BNT162b2 (BioNTech/Pfizer) vaccines, suggesting a substantial induction of B cell memory responses. Noteworthy, three obese previously seropositive individuals failed to mount a booster response upon vaccination, warranting further studies in this population. Immunization of naïve individuals with two doses of the CoronaVac vaccine or one dose of the BNT162b2 vaccine elicited similar levels of nAbs compared to seropositive individuals 4.2 to 13.3 months post-infection with SARS-CoV-2. Thus, this preliminary evidence suggests that both, seropositive and naïve individuals, require two doses of CoronaVac to ensure the induction of robust nAb titers.

## MAIN TEXT

The durability of circulating neutralizing antibody (nAb) responses to severe acute respiratory syndrome coronavirus 2 (SARS-CoV-2) infection or vaccination has become a central question during the current pandemic to determine correlates of protection against disease. Current evidence shows that SARS-CoV-2 spike-specific antibodies decline over time but remain detectable up to 8 months post-symptoms onset^1^. However, additional longitudinal data are needed to characterize the medium- and long-term protective antibody dynamics, starting from the acute phase of disease of patients with mild and moderate/severe outcome, and to determine their nAb memory responses upon immunization with different vaccines currently in use.

We enrolled 74 individuals (overall mean age 44 years [range 14 to 83, >60 23%]), of whom 37 were outpatient (mild disease, mean age 37 years [range 14 to 66]) and 37 were hospitalized (moderate and severe disease, mean age 51 years [range 16 to 83]) with a confirmed SARS-CoV-2 quantitative RT-PCR test (Suppl. Table 1). These individuals were followed longitudinally to determine nAb response for up to one year from the onset of symptoms (demographic and baseline characteristics of the patients are summarized in Suppl. Table 1; samples were collected between 2 to 414 days after the onset of symptoms).

Regardless of disease severity, infected individuals developed robust nAb responses during the first month. These responses declined over time but were sustained for up to 12 months (Fig. 1A, B), as determined with a microneutralization assay based on a recombinant vesicular stomatitis virus carrying a SARS-CoV-2 spike protein that showed strong correlation (Pearson’s r = 0.86, R2 = 0.75, P< 0.001) with authentic SARS-CoV-2 microneutralization (Fig. S1). We performed kinetic analyses with samples from 41 individuals that were sampled weekly during the first month from symptom onset (Fig. 1A and Fig. S2A-B). In agreement with previous reports, hospitalized individuals had significantly higher neutralization titers as compared to outpatients (Fig. S2C), with peak average nAb responses at day 23 and at day 27 post-symptom onset, respectively (Fig. 1A and Fig. S2A-B). We included longitudinal samples for all participants and performed a nAb titer time decay analysis starting from the respective peak average responses. Fitting our nAb data to a one-phase decay model, the initial decay half time was 42 days (95% CI : 2.21 to 362.4) for outpatients and 84 days (95% CI : 1.7 to indeterminate) for hospitalized individuals, and when we used a continuous decay fit, the half time was 225 (95% CI : 121 to 1,648) and 195 (95% CI : 120 to 535) days for these groups, respectively (Fig. 1B). None of the individuals in the study had evidence of re-infections. Hence, albeit the nAb titers remained higher in hospitalized patients than in the outpatients, both study groups showed long-lasting responses of circulating antibodies after natural infection.

**Figure 1.**
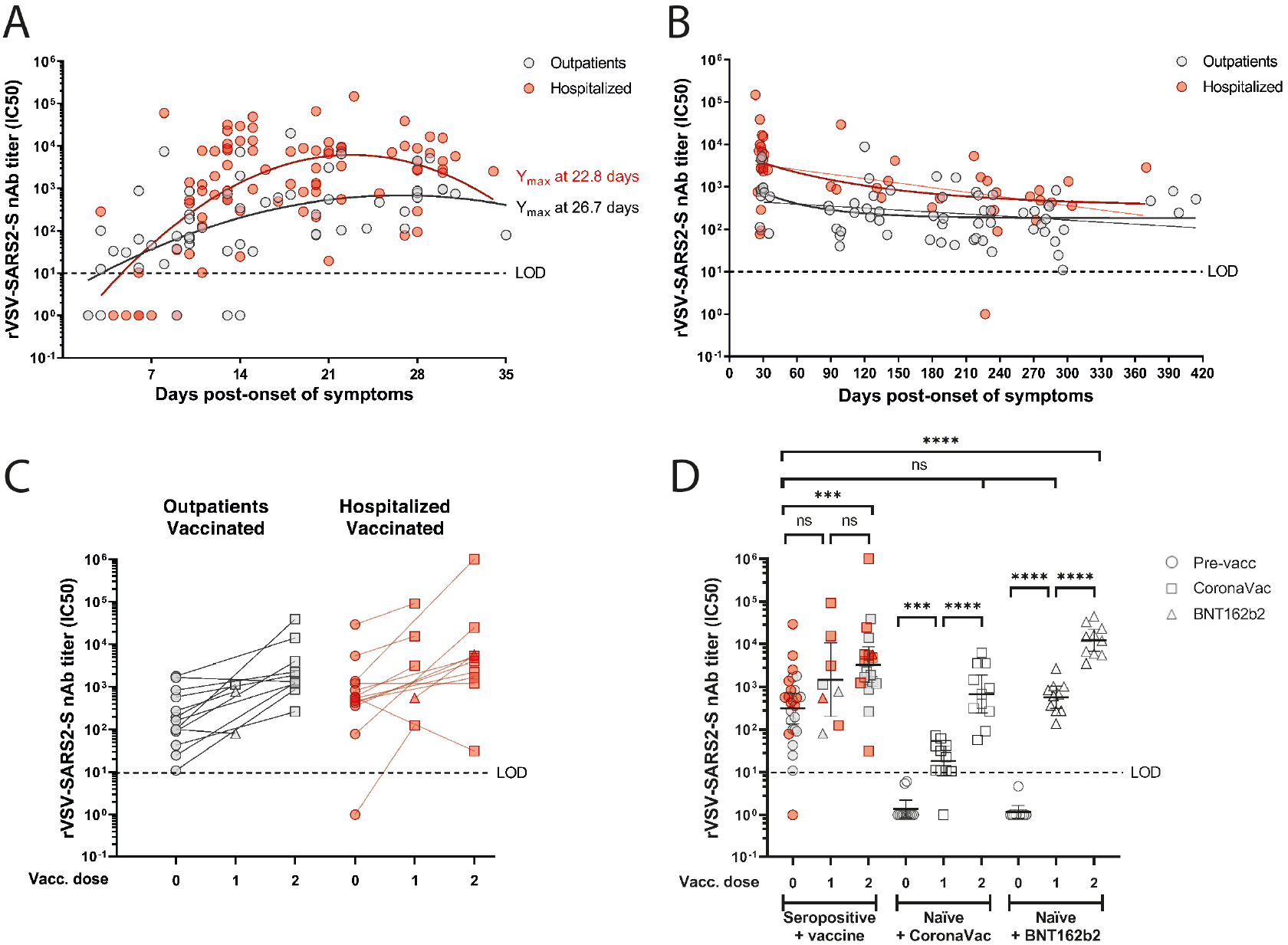
Neutralizing antibody responses to SARS-CoV-2 in seropositive and naïve individuals before and after CoronaVac or BNT162b2 vaccination. (Panels A to D) The half-maximum inhibitory concentration (IC50) of sera was determined by microneutralization assay of recombinant vesicular stomatitis virus carrying SARS-CoV-2 spike protein (rVSV-SARS2-S). (Panel A) Neutralizing antibody (nAb) titers (IC50) from 15 outpatients (57 samples; grey circles) and 26 hospitalized (84 samples; red circles) at 2 to 36 days post-symptom onset. Second order polynomial (quadratic) curve fitting was used to establish the days at which peak titers occurred (Ymax). (Panel B) Longitudinal nAb titers from 36 outpatients (66 samples) and 31 hospitalized (44 samples) taken from day 27 (outpatients) or day 23 (hospitalized) until day 414 post-symptom onset. One-phase decay fit is indicated as a bold line, while continuous decay fit is shown with the thinner line in red and gray for the corresponding patient group. (Panel C) nAb titers from 13 outpatient (26 samples) or 14 hospitalized (28 samples) individuals immunized with one or two doses of CoronaVac (24 participants) or one or two doses of BNT162b2 (3 participants) vaccines. (Panel D) nAb titers from naïve individuals after the first and second dose of CoronaVac (11 participants) or BNT162b2 (10 participants) vaccines, compared to seropositive individuals who were not vaccinated (26 participants) or received one dose (8 samples) or two doses (20 samples) of the indicated vaccines. Geometric means with 95% confidence intervals are shown. Circles, non-vaccinated; squares, vaccinated with CoronaVac; triangles, vaccinated with BNT162b2. Dashed line indicates the limit of detection (LOD) of the microneutralization assay. Statistics were performed using unpaired two-tailed Mann-Whitney test. ***P<0.001; ****P<0.0001; ns, non-significant.

Twenty-seven of the previously seropositive individuals (mean age 45 years [range 17 to 83]) included in our longitudinally cohort study were immunized during the study period. Thus, we analyzed the nAb response in these previously infected individuals after immunization with the two main vaccines currently being used in Chile; the CoronaVac (Sinovac) vaccine based on inactivated virus or the BNT162b2 (BioNTech/Pfizer) vaccine based on spike protein-encoding messenger RNA. We compared them to nAbs titers in healthy SARS-CoV-2 naïve (seronegative) individuals immunized with two doses of either vaccine (CoronaVac, 11 participants, mean age 34 years [range 21 to 47] or BNT162b2, 10 participants, mean age 37 years [range 23 to 53]; Suppl. Table 1). The seropositive individuals were vaccinated between 4.2 to 13.3 months (average 9.9 months) after the onset of symptoms for both, the outpatient (13 participants) and hospitalized groups (14 participants; see arrows in Fig. S3). The average increase in the nAb titers was four times for outpatients (pre-vaccine mean titer = 188.9 [range 98.7 to 273.6], first dose mean titer = 666.3 [range 80.9 to 1,143]) and three times for hospitalized individuals (pre-vaccine mean titer = 8,836 [range 1.0 to 29,389], first dose mean titer = 27,562 [range = 124.7 to 91,571]) after one dose (Fig. 1C). After the second dose, the average increase was 13 times (pre-vaccine mean titer = 528.7 [range 10.9 to 1,791], mean titer = 6,597 [range 261.6 to 38,912) and 179 times (pre-vaccine mean titer = 649.1 [range 78.0 to 1,360], mean titer = 116,012 [range 31.0 to 1,000,000]), respectively (Fig. 1C). Except for four cases, all participants showed an increase in the nAb titer after receiving one or two doses of the vaccines, suggesting a significant induction of B cell memory response at 4.2 to 13.3 months after onset of symptoms. One corresponded to an outpatient (Fig. S3C, dark yellow patient) who had been vaccinated two days earlier and may therefore not have had sufficient time to mount a booster response. The other three participants (age range 29 to 63 years) were obese outpatient (Fig. S3C, light green patient) or hospitalized (Fig. S3D, grey and cyan patients) individuals for whom we only had a previous sample 5.6 to 10.7 months prior to vaccination (Fig. S3C, D), and hence no clear conclusions can be drawn about the trajectory of their nAb titers. Noteworthy, one of these participants had a marked decreased nAb titer after two doses of the vaccine (IC50 563.9 to 31.0; Fig. 1C). Due to the high prevalence of obesity in severe coronavirus disease 2019 (COVID-19)^2^, further studies to monitor the induction of nAbs after vaccination in this population are required.

The induction of nAbs in seropositive individuals vaccinated with one dose as compared to the naïve individuals vaccinated with BNT162b2 was on average 18 times higher (previously infected first dose mean titer = 14,099 [range 80.91 to 91,571], naïve first dose mean titer = 791.0 [range 137.0 to 2,663]). When comparing them with naïve individuals vaccinated with one dose of CoronaVac, their nAb titers were on average 492 times higher (naïve first dose mean titer = 28.64 [range 1.0 to 71.6]). Importantly, we did not detect a statistical difference between nAb titers of seropositive individuals 0.9 to 10.1 months post-infection (mean titer = 1,860 [range 1.0 to 29,389]) with those of naïve individuals vaccinated with two doses of CoronaVac (mean titer = 1,595 [range 56.9 to 6,262]), and higher titers were observed in those individuals with two doses of BNT162b2 (mean titer = 16,648 [range 3,500 to 44,719]), suggesting the induction of robust nAbs responses in naïve individuals with both vaccines (Fig. 1D). Immunization of previously infected individuals with CoronaVac (24 participants), showed no significant differences in nAb titers after one dose or when comparing the first and second dose. Significantly increased nAb titers were only observed after both doses (P<0.01) of this vaccine (Fig. 1D), differing from vaccination of seropositive or naïve individuals with mRNA and adenovirus-based vaccines that induce high nAb titers after the first dose^3,4^.

We found long lasting nAb titers that persist for over 12 months after the onset of symptoms in both, outpatient and hospitalized individuals. Neutralizing activity in seropositive individuals was boosted significantly after two doses of the CoronaVac or BNT162b2 vaccines, regardless of the time interval since the onset of COVID-19 symptoms, suggesting that infection induces a robust B-cell memory response. The correlates of protection against SARS-CoV-2 are currently unknown. However, current evidence of re-infections remains limited, and they appear to be infrequent, suggesting that natural infection provides significant protection against COVID-19 in most individuals^5^. While this is a small cohort study, our results indicate that natural infection induces long-lasting nAb responses that can be significantly boosted through vaccination, and that immunization of naïve individuals with two doses of the CoronaVac vaccine or one dose of the BNT162b2 vaccine elicit similar levels of nAbs compared to seropositive individuals 4.2 to 13.3 months post-infection with SARS-CoV-2. Our preliminary evidence suggests that both, seropositive and naïve individuals, require two doses of CoronaVac to generate a robust induction of nAb titers. Further studies to determine the long-term duration of vaccine-induced nAbs against SARS-CoV-2 are warranted.

## Data Availability

The data supporting the findings of this study are available within the article and its supplementary material. The raw data that support the findings of this study are available on request from the corresponding author.

## Author Contribution

NAM and TGS collected and analyzed data, made figures and tables, interpreted data, and wrote the paper. CPR, EFS, JL, MJA, LIA, EP, and SS processed samples, performed experiments, analyzed data, and revised the paper. GV, ES, CG, AR, recruited patients, collected clinical metadata, and revised the paper. RJ, KC, DH, MED, generated rVSV viral stocks, analyzed data and revised the paper. FK analyzed serological data, advised on data interpretation, and provided funding for the study and revised the paper. NT designed the study, collected, analyzed, and interpreted data, provided funding for the study, and wrote the paper. RAM conceived the longitudinal cohort design, recruited patients; collected, analyzed, and interpreted data, provided funding for the study, and wrote the paper.

## SUPPLEMENTARY APPENDIX

### Supplemental Material and Methods

#### Study population and clinical metadata

74 individuals with a confirmed diagnosis for SARS-CoV-2 infection were recruited between March 5 and October 22, 2020. Patient clinical and epidemiological data, along with their clinical specimens were collected after informed written consent was obtained under protocols 16-066 and 200829003 which were reviewed and approved by the Scientific Ethics Committee at Pontificia Universidad Católica de Chile (PUC). The analysis were performed considering two major groups of individuals, hospitalized and outpatients: **Hospitalized** individuals were either severe patients, defined as those who developed pneumonia with one of the following three conditions: (1) acute respiratory failure that required invasive mechanical ventilation or a high-flow nasal cannula (HFNC) with prone position, (2) septic shock or (3) multiple organ dysfunction; moderate cases consisted of inpatients with pneumonia without these conditions. **Outpatients** were individuals that had mild symptoms of COVID-19 but did not meet the above-mentioned criteria. Peripheral blood samples, nasopharyngeal swabs and sputum samples were collected between 2 and 414 days after the onset of symptoms. Healthy individuals were recruited as controls and received the two doses of the CoronaVac (Sinovac Life Sciences Co., LTD, Beijing, China) or the BNT162b2 (Pfizer Manufacturing Belgium NV, Puurs, Belgium) vaccines. Samples were collected between 20-30 days after first dose but prior to the second dose and 13-19 days after second dose. Demographic data for all patients and controls, obtained by a clinical questionnaire, are shown in Supplementary Table 1.

#### Plasma and serum collection

Peripheral blood was collected in both plasma separating (EDTA/purple top) and serum separating (red top) tubes and was processed by centrifugation at 3000 rpm for 5 min. Plasma and serum samples were aliquoted and stored at −80°C. Serum samples were heated at 56°C for 1h before use to eliminate the risk of any potential residual virus.

#### SARS-CoV-2 spike ELISA

Overnight, 96-well plates (Immulon 4 HBX; Thermo Fisher Scientific) were coated at 4°C with 50 µL per well of a 2 mg/mL solution recombinant SARS-CoV-2 spike protein suspended in phosphate-buffered saline (PBS, Gibco), as previously described^1,2^. The next morning, the coating solution was removed and 100 µL per well of 3% non-fat milk prepared in PBS with 0.1% Tween 20 (PBST) was added to the plates as a blocking solution and incubated at room temperature for 1h. Serial dilutions of serum and antibody samples were prepared in 1% non-fat milk prepared in PBST. The blocking solution was removed and 100 µL of each serial dilution was added to the plates for 2 h at room temperature. Plates were washed three times with 250 µL per well of 0.1% PBST. Next, a 1:3,000 dilution of goat anti-human IgG–horseradish peroxidase (HRP) conjugated secondary antibody (Thermo Fisher Scientific) was prepared in 0.1% PBST and 100 µL of this secondary antibody was added to each well for 1 h. Plates were again washed three times with 0.1% PBST. Once completely dry, 100 µL SIGMAFAST OPD (o-phenylenediamine dihydrochloride; Sigma–Aldrich) solution was added to each well. This substrate was left on the plates for 10 min and then the reaction was stopped by the addition of 50 µL per well of 3 M hydrochloric acid. The optical density at 490 nm (OD490) was measured using a Synergy 4 (BioTek) plate reader. The background value was set at an OD490 of 0.11 and the area under the curve (AUC) was calculated. AUC values below 1 were assigned a value of 0.5 for plotting and calculation purposes. In some cases, end-point titers were calculated, with the end-point titer being the last dilution before reactivity dropped below an OD490 of <0.11. CR3022, a human monoclonal antibody reactive to the RBD of both SARS-CoV-1 and SARS-CoV-2^3,4^, was used as control.

#### SARS-CoV-2 microneutralization assay

Vero E6 cells were seeded at a density of 20,000 cells per well in a 96-well cell culture plate in complete Dulbecco’s Modified Eagle Medium (cDMEM). The following day, heat-inactivated serum samples (dilution of 1:10) were serially diluted threefold in 1× MEM (10% 10× minimal essential medium (Gibco), 2 mM L-glutamine, 0.1% sodium bicarbonate (wt/vol; Gibco), 10 mM 4-(2-hydroxyethyl)-1-piperazineethanesulfonic acid (HEPES; Gibco), 100 U/mL penicillin, 100 mg/mL streptomycin (Gibco) and 0.2% bovine serum albumin (MP Biomedicals). As previously described^5^, the authentic SARS-CoV-2 (USA-WA1/2020; GenBank: MT020880) was diluted to a concentration of 100 TCID50 (50% tissue culture infectious dose) in 1× MEM. Then, 80 µL of each serum dilution and 80 µL of the virus dilution were added to a 96-well cell culture plate and allowed to incubate for 1h at room temperature. cDMEM was removed from Vero E6 cells and 120 µL of the virus–serum mixture was added to the cells. Then, the cells were incubated at 37°C for 1h. After the 1 h incubation, the virus–serum mixture was removed from the cells and 100 µL of each corresponding serum dilution and 100 µL of 1× MEM containing 1% fetal bovine serum (FBS, Corning) was added to the cells. The cells were incubated for 48 h at 37°C and then fixed with 10% paraformaldehyde (Polysciences) for 24 h at 4°C. Following fixation, the paraformaldehyde was removed, and the cells were washed with 200 µL PBS. The cells were then permeabilized by the addition of 150 µL PBS containing 0.1% Triton X-100 for 15 min at room temperature. The plates were then washed three times with PBS containing PBST and blocked in [3% milk (American Bio) in PBST] solution for 1 h at room temperature. After blocking, 100 µL of mAb 1C7 (anti-SARS nucleoprotein antibody generated in-house) at a dilution of 1:1,000 was added to all wells and the plates were allowed to incubate for 1h at room temperature. The plates were then washed three times with PBST before the addition of goat anti-mouse IgG–HRP (Rockland Immunochemicals), diluted 1:3,000 in blocking solution for 1 h at room temperature. Plates were then washed three times with PBST, and SIGMAFAST OPD (Sigma–Aldrich) was added. After a 10 min incubation at room temperature, the reaction was stopped by adding 50 µL 3 M hydrochloric acid to the mixture. The OD490 was measured on a Synergy 4 plate reader (BioTek). A cut-off value of the average of the optical density values of blank wells plus three standard deviations was established for each plate and used to calculate the microneutralization titer. Microneutralization assays were performed in a facility with a biosafety level of 3.

#### rVSV SARS-CoV-2 spike protein (rVSV-SARS2-S) microneutralization assay

To determine the nAb titers of patient sera, the replication-competent recombinant vesicular stomatitis virus previously described by Dieterle et al. (2020) was used, in whose genome the native glycoprotein gene has been replaced by that encoding the spike glycoprotein of the SARS-CoV-2 and whose genome further encodes an enhanced green fluorescent protein (eGFP)^6^. This system has the advantage of generating high viral titers (~10^7^), an easy score of cell infection by GFP fluorescence, correlates of convalescent serum-mediated neutralization with that of authentic SARS-CoV-2, enters cells through pathways of SARS-CoV-2, and does not require high biosafety containment infrastructure for manipulation^5^. Briefly, Vero E6 cells (ATCC) grown in 1X MEM supplemented with 10% FBS (Thermo Scientific) were transfected with plasmid pCEP4-myc-ACE2 (Addgene catalog # 141185) and stable clones selected in presence of hygromycin (400 µg/mL) based on the presence of high levels of hACE2 at the plasma membrane. To assay nAb titers, serial dilutions of serum samples were mixed with rVSV-SARS2-S and incubated for 1 h at 37°C. The serum-virus was subsequently added to Vero E6 hACE2 cells seeded the day before in optical bottom 96-well plates (Thermo Scientific) at 80% confluence and adsorbed for 2 h at 37°C. Next, the mixture was replaced by culture media and infection allowed to proceed for 20 h at 37°C, 5% CO2 and 80% humidity. Next, the cells were fixed with 4% formaldehyde (Thermo Scientific), washed with PBS, stained in with 4′,6-diamidino-2-phenylindole (DAPI) 300 nM (Thermo Scientific) and stored in PBS at 4°C until acquiring fluorescence data. Viral infectivity was either quantified by automated enumeration of GFP-positive cells (normalizing against cells stained with DAPI) using a Cytation5 automated fluorescence microscope (BioTek) and segmentation algorithms applied from the ImageJ program. Alternatively, total GFP fluorescence per well was acquired using the Cytation5 fluorescence lector (wavelength for DAPI 360 nm for absorption, 460 nm for emission and for GFP, 485 nm for absorption, 526 nm for emission) and normalized against DAPI fluorescence. The half-maximum inhibitory concentration (IC50) of the sera, calculated using non-linear regression analysis and curve fitting using second-order polynomial (quadratic), one phase decay and linear regression models (using log10 IC50 transformed data) were done with GraphPad Prism 5 software.

#### Computational and statistical analysis

Categorical variables were expressed as numbers or percentages. Association between categorical variables was examined with Chi-squared or Fisher’s exact test. Continuous variables were expressed in mean, geometric mean and range and compared with unpaired two-tailed Mann-Whitney test. Correlation was evaluated calculating the Pearson correlation coefficient. GraphPad Prism 8 was used for statistical analysis. * P<0.05; ** P<0.01; *** P<0.001; **** P<0.0001.

---

**Supplementary Table 1.** Demographic and baseline characteristics of COVID-19 patients and vaccinated controls.

|  | Outpatients<br>(37<br>participants) | Hospitalized<br>(37<br>participants) | P value<br>(Outpatients<br>vs.<br>Hospitalized<br>) | CoronaVac<br>(11<br>participants) | BNT162b2<br>(10<br>participants) | P value<br>(CoronaVac<br>vs.<br>BNT162b2) |
| --- | --- | --- | --- | --- | --- | --- |
| **Characteristics** |  |  |  |  |  |  |
| Male, n (%) | 17 (45.9) | 25 (67.6) | 0.0998 | 4 (36.4) | 2 (20) | 0.6351 |
| Age, mean (range) | 37 (14-66) | 51 (16-83) | **0.0004** | 34 (21-47) | 37 (23-53) | 0.6981 |
| >60 years, n (%) | 5 (13.5) | 12 (32.4) | 0.0956 | 0 | 0 | - |
| **Symptoms** |  |  |  |  |  |  |
| **Respiratory** |  |  |  |  |  |  |
| Cough, n (%) | 27 (73) | 31 (83.8) | 0.3975 |  |  |  |
| Dyspnea, n (%) | 6 (16.2) | 19 (51.4) | **0.0028** |  |  |  |
| Odynophagia, n (%) | 21 (56.8) | 6 (16.2) | **0.0006** |  |  |  |
| Chest discomfort, n (%) | 3 (8.1) | 5 (13.5) | 0.7106 |  |  |  |
| **Constitutional** |  |  |  |  |  |  |
| Fever, n (%) | 22 (59.5) | 31 (83.8) | **0.0377** |  |  |  |
| Headache, n (%) | 32 (86.5) | 14 (37.8) | **< 0.0001** |  |  |  |
| Myalgia, n (%) | 25 (67.6) | 18 (48.6) | 0.157 |  |  |  |
| Severe fatigue, n (%) | 0 | 20 (54.1) | **< 0.0001** |  |  |  |
| Altered mental status, n (%) | 0 | 3 (8.1) | 0.2397 |  |  |  |
| **Gastrointestinal** |  |  |  |  |  |  |
| Diarrhea, n (%) | 12 (32.4) | 10 (27) | 0.7997 |  |  |  |
| Nausea/Vomiting, n (%) | 6 (16.2) | 9 (24.3) | 0.5642 |  |  |  |
| **Sensorial** |  |  |  |  |  |  |
| Ageusia, n (%) | 18 (48.6) | 5 (13.5) | **0.0022** |  |  |  |
| Anosmia, n (%) | 24 (64.9) | 8 (21.6) | **0.0004** |  |  |  |
| **Comorbidities or conditions** |  |  |  |  |  |  |
| Obesity (BMI ≥ 30), n (%) | 5 (13.5) | 14 (37.8) | **0.0317** | 2 (18.2) | 4 (40) | 0.3615 |
| Hypertension, n (%) | 3 (8.1) | 13 (35.1) | **0.0095** | 0 | 2 (20) | 0.2143 |
| Metabolic conditions*, n (%) | 4 (10.8) | 12 (32.4) | **0.0459** | 1 (9.1) | 1 (10) | 1 |
| Hyperlipidemia, n (%) | 4 (10.8) | 7 (18.9) | 0.5151 | 0 | 2 (20) | 0.2143 |
| Cardiovascular disease, n (%) | 0 | 3 (8.1) | 0.2397 | 0 | 0 | - |
| Chronic pulmonary disease, n (%) | 4 (10.8) | 3 (8.1) | 1 | 0 | 0 | - |
| Asthma, n (%) | 6 (16.2) | 2 (5.4) | 0.2611 | 1 (9.1) | 1 (10) | 1 |
| Rheumatologic disease, n (%) | 0 | 3 (8.1) | 0.2397 | 0 | 1 (10) | 0.4762 |
| Immunocompromised, n (%) | 0 | 5 (13.5) | 0.0541 | 0 | 1 (10) | 0.4762 |
| Allergy**, n (%) | 16 (43.2) | 6 (16.2) | **0.0209** | 7 (63.6) | 2 (20) | 0.0805 |
| Neurologic disease, n (%) | 0 | 4 (10.8) | 0.1148 | 0 | 0 | - |
| Smoker, n (%) | 8 (21.6) | 9 (24.3) | 1 | 1 (9.1) | 3 (30) | 0.3108 |

Abbreviation: BMI, Body mass index. \*Metabolic conditions include insulin resistance, prediabetes, type 1/2 diabetes, non-alcoholic steatohepatitis and obstructive sleep apnea; \*\*Allergy considered self-reported allergic rhinitis (by seasonal, perennial/year-round, or episodic allergens) and food allergy.

**Supplementary Figure 1.**
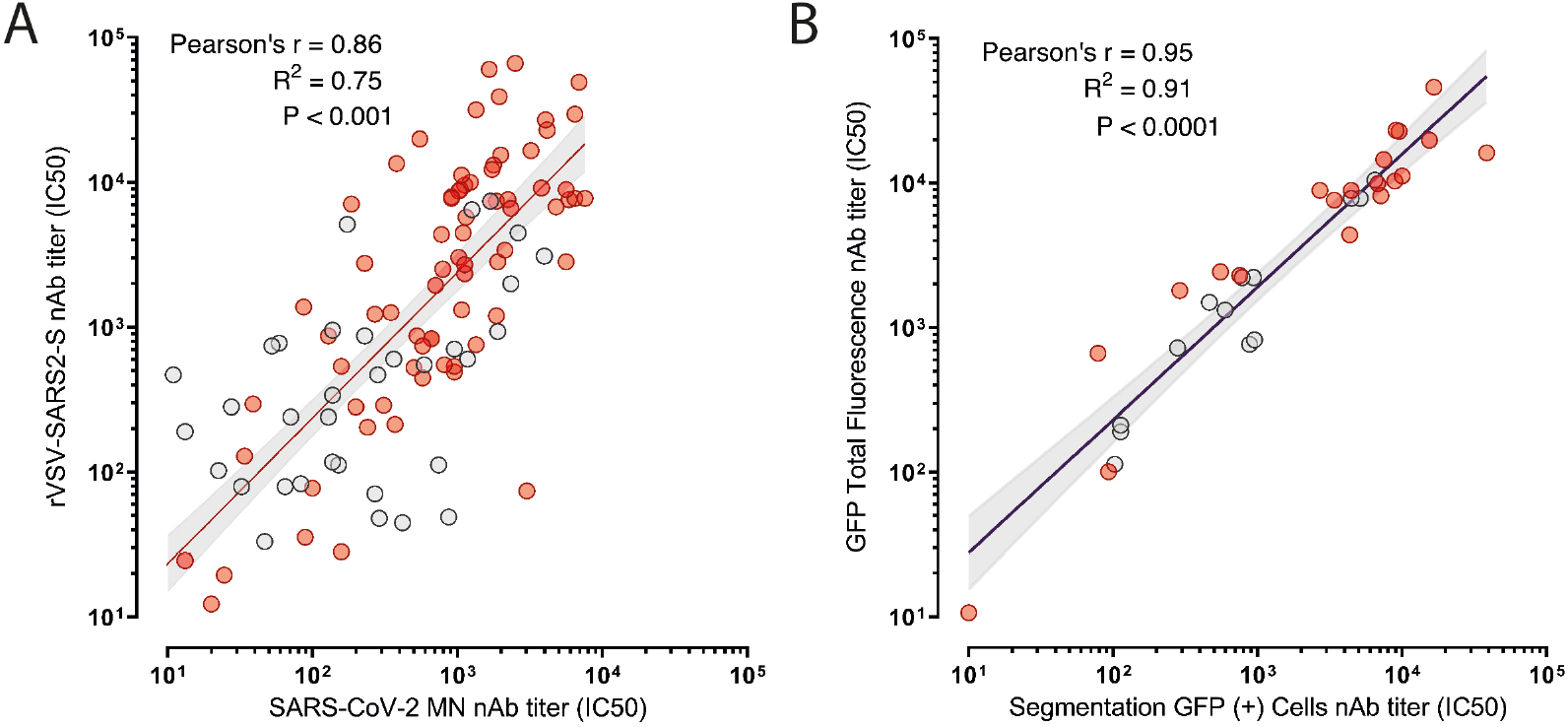
Correlation of the neutralizing activity of convalescent sera in neutralization assays with rVSV-SARS2-S and to the authentic SARS-CoV-2 virus. (Panel A) Linear regression and Pearson correlation analysis comparing neutralizing activities of seropositive sera with authentic SARS-CoV-2 and rVSV-SARS2-S (122 samples; outpatients, grey circles; hospitalized, red circles). (Panel B) Linear regression and Pearson correlation analysis from two different quantification methods to measure neutralizing activity against rVSV-SARS2-S; GFP (+) cell counting by using ImageJ segmentation analysis and total GFP measurement normalized to DAPI counterstaining (33 samples).

**Supplementary Figure 2.**
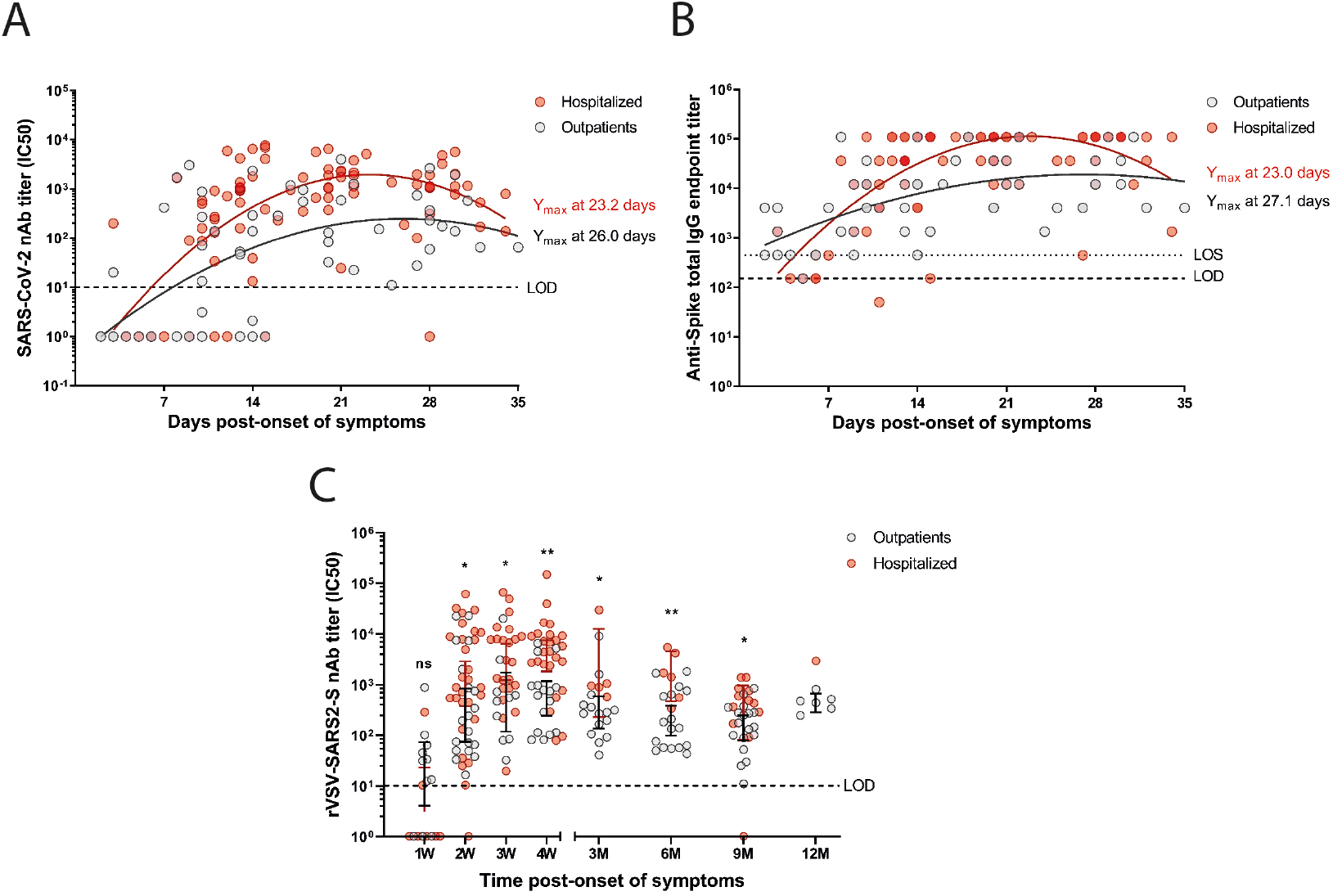
Longitudinal dynamics of humoral responses to the spike protein of SARS-CoV-2 from outpatient and hospitalized seropositive individuals. (Panel A) nAbs titers against authentic SARS-CoV-2 of sera obtained from 41 seropositive individuals (142 samples) during 35 days since symptom onset. (Panel B) Anti-Spike total IgG endpoint titer of serially diluted sera from 41 seropositive individuals (145 samples) during 36 days since symptom onset determined by an enzyme-linked immunosorbent assay (ELISA). Second order polynomial (quadratic) curve fitting was used to establish the days at which peak titers occurred (Ymax). (Panel C) Neutralizing antibody (nAb) titers (IC50) obtained using rVSV-SARS2-S microneutralization assay for 37 outpatients (111 samples; grey circles) and 37 hospitalized (108 samples; red circles) grouped by weeks (W) or months (M) post-symptom onset. The bars indicate geometric mean titers with 95% confidence intervals. Dotted line represents the limit of sensitivity (LOS) of ELISA. Dashed line represents the limit of detection (LOD) of each assay. Statistics were calculated at the indicated time points between nAb titers of outpatient and hospitalized using the unpaired two-tailed Mann-Whitney test (*P<0.05; **P<0.01).

**Supplementary Figure 3.**
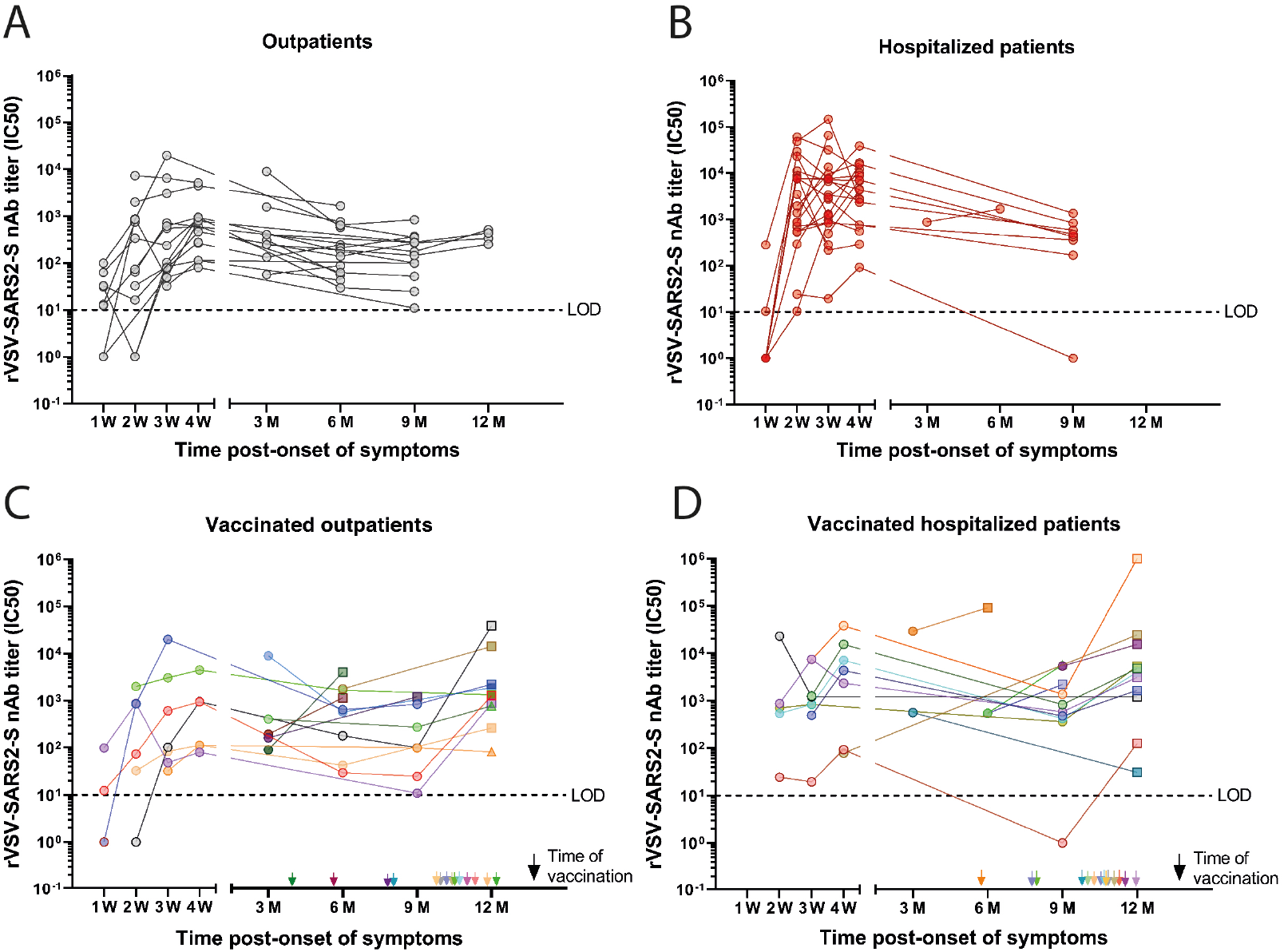
Longitudinal responses of seropositive individuals with or without CoronaVac and BNT162b2 vaccination. Longitudinal neutralizing antibody (nAb) titers (IC50) obtained using an rVSV-SARS2-S microneutralization assay for seropositive outpatients (Panel A; 24 participants), hospitalized patients (Panel B; 25 participants), vaccinated seropositive outpatients (Panel C; 13 participants) or vaccinated hospitalized patients (Panel D; 14 participants) at different time points grouped by weeks (W) or months (M) post-symptom onset (serum samples from: 3M = 46-135 days; 6M = 136-225 days; 9M = 226-315 days and 12M = 316-414 days). The arrows indicate time of vaccination post-onset of symptoms. Circles, non-vaccinated; squares, vaccinated with CoronaVac; triangles, vaccinated with BNT162b2. Dashed line indicates the limit of detection (LOD) of the microneutralization assay.

